# Chronic Hemodialysis Patients have better outcomes with COVID-19 - a retrospective cohort study

**DOI:** 10.1101/2020.07.22.20159202

**Authors:** Ashutossh Naaraayan, Abhishek Nimkar, Amrah Hasan, Sushil Pant, Momcilo Durdevic, Henrik Elenius, Corina Nava Suarez, Prasanta Basak, Kameswari Lakshmi, Michael Mandel, Stephen Jesmajian

## Abstract

**Introduction:** Several comorbid conditions, have been identified as risk factors in patients with COVID-19. However, there is a dearth of data describing the impact of COVID-19 infection in patients with end-stage renal disease on hemodialysis (ESRD-HD).

**Methods:** This retrospective case series analyzed 362 adult patients consecutively hospitalized with confirmed COVID-19 illness between March 12, 2020 and May 13, 2020, at a teaching hospital in the New York City metropolitan area. Primary outcome was severe pneumonia as defined by the World Health Organization. Secondary outcomes were: 1) the Combined Outcome of Acute respiratory distress syndrome or in-hospital Death (COAD), and 2) the need for High-levels of Oxygen supplementation (HiO2).

**Results:** Patients with ESRD-HD had lower odds for poor outcomes including severe pneumonia [Odds Ratio (OR) 0.4, Confidence Interval (CI) (0.2–0.9) p=.04], HiO2 [OR 0.3, CI (0.1– 0.8) p=.02] and COAD [OR 0.4, CI (0.2–1.05) p=.06], when compared to patients without ESRD. In contrast, higher odds for severe pneumonia, COAD and HiO2 were seen with advancing age. African-Americans were over-represented in the hospitalized patient cohort, when compared to their representation in the community (35% vs 18%). Hispanics had higher odds for severe-illness and HiO2 when compared to Caucasians.

**Conclusions:** Patients with ESRD-HD had a milder course of illness with a lower likelihood of severe pneumonia and a lesser need for aggressive oxygen supplementation when compared to patients not on chronic dialysis. This “protective effect” might have a pathophysiologic basis and needs to be further explored.

## Introduction

At the time of writing, fourteen million cases of coronavirus disease 2019 (COVID-19) caused by severe acute respiratory syndrome coronavirus-2 (SARS-CoV-2) have been described worldwide, resulting in 600 000 deaths.^1^ COVID-19 was declared a pandemic by the World Health Organization (WHO) on March 11, 2020 and the New York Metropolitan region was at its epicenter through March and April, 2020.^2^

So far, laboratory confirmed cases have been documented from 188 countries on six continents, with the US reporting the highest number of cases and deaths.^1^

It is crucial to identify the demographic and comorbid factors that impact illness severity among hospitalized COVID-19 patients. Several comorbid conditions such as hypertension, diabetes, chronic obstructive pulmonary disease and atherosclerotic cardiovascular diseases, have been identified as risk factors in patients with COVID-19.^3,4^ However, there is a dearth of data describing the impact of COVID-19 infection in patients with end-stage renal disease on hemodialysis (ESRD-HD).

Being on the frontlines at one of the earliest epicenters in the US,^5^ we observed that patients with ESRD-HD were having a milder course of illness compared to patients not on hemodialysis (HD). Here we present data analyzing the impact of patient characteristics on illness severity among hospitalized COVID-19 patients, while specifically investigating the effect of ESRD-HD on outcomes.

## Methods

### Data source

This retrospective case series includes patients consecutively hospitalized with confirmed COVID-19 illness between March 10, 2020 and May 13, 2020, at a 242-bed teaching community hospital in the NYC metropolitan region. Cases were confirmed through positive result for SARS-CoV-2 virus by reverse-transcriptase-polymerase-chain-reaction testing of nasopharyngeal swab specimen. Data was manually abstracted from electronic health records by the authors, and included demographics, comorbid conditions and clinical data (respiratory rate, oxygen saturations, arterial blood gas tests when available, type/level of oxygen supplementation). Three authors (AN, AN and SP) independently reviewed the data for accuracy.

Hospital course was followed up until June 15, 2020. Details on the criteria for hospitalization are provided in the Supplementary Materials. Of the 377 patients admitted during the study period, those who were transferred to another facility for tertiary level care (n=5), those who left against medical advice (n=2) and those who were still receiving care at our hospital (n=8) were excluded from the analysis. After said exclusions, final analysis included 362 patients.

### Definition of patient characteristics

Comorbidities derived from the patients or nursing home transfer forms were abstracted from physician documentation on the electronic health records. Patients were classified into three categories based on the presence or absence of kidney disease; i) no chronic kidney disease, ii) presence of chronic kidney disease, and iii) presence of end-stage

renal disease on hemodialysis (ESRD-HD). Outcomes were analyzed in patients with chronic kidney disease versus those without kidney disease and patients with ESRD-HD versus those without kidney disease. Cardiac disease was defined as chronic heart conditions including; previous myocardial infarction, cardiac arrhythmias, congestive heart failure, presence of pacemaker or defibrillator device and previous coronary artery bypass grafting or percutaneous coronary intervention. Body mass index >=30 kg/m^2^ was used to identify obesity. Race/ethnicity was classified into one of the four categories: Caucasian, African-American, Hispanic ethnicity regardless of race, and others.

Severe disease was defined as clinical pneumonia with a respiratory rate >30 breaths/minute or oxygen saturation <90% on room air, per the WHO guidelines.^6^ Acute respiratory distress syndrome (ARDS) was defined as the ratio of arterial oxygen partial pressure to fractional inspired oxygen of <=300 on a positive end-expiratory pressure >= 5cm of water, as per the Berlin guidelines.^7^ Mortality was defined as in-hospital death. Need for aggressive oxygen supplementation with high-flow nasal canula, non-invasive positive pressure ventilation or mechanical ventilation was defined as High-oxygen support (HiO2).

### Outcome measures and statistical analysis

The study was done according to the STROBE guidelines for observational studies.^8^ We computed median with inter-quartile range, frequency, and percentages as our descriptive variables. Differences in median were calculated using the Mann-Whitney test. Differences in percentage were assessed using the chi-squared test. Our primary outcome was development of severe pneumonia as defined by the WHO. Our secondary outcomes were: 1) the need for High-oxygen support (HiO2), and 2) the Combined Outcome of a diagnosis of ARDS or in-hospital Death (COAD).

Logistic regression analysis by univariate and age-adjusted models, were performed for patient characteristics. Subsequently, to determine independent association between these characteristics and outcomes, multivariable logistic regression models were used to estimate odds ratios adjusted for clinical covariates. Multivariable analysis was performed using the least absolute shrinkage and selection operator (Lasso) inference, a standard model building approach.^9^ Lasso is most useful for sparse high-dimensional models such as ours. Double selection was used for estimation as this inference method provides the most stable results. Stata version 16.0 was used for analysis (Stata Corp, Houston, Texas). Age, sex, hypertension, diabetes and cardiac disease were always included for adjustment in this model. Lasso was asked to select variables in the model from the following specified covariates (race, chronic obstructive pulmonary disease, renal disease and obesity). Each variable of interest was fitted separately for multivariable analysis. When one of these covariates was the variable of interest, it was not specified to Lasso for model selection. Overfitting and collinearity were resolved by using Lasso and defining variables as stated above. Two-sided p<0.05 was considered statistically significant.

### Ethics

The study was approved by the departmental research review committee with a waiver of informed consent due to its retrospective design [Approval number 20.5.02].

## Results

Among the 362 patients, the median age was 71 years (59–82), 55.3% were men, 35.4% were African-American and 232 (63%) had severe disease (Table 1). There were no deaths among patients without severe disease. Length of hospital-stay was longer among patients with severe disease. Patients with severe disease were more likely to be Hispanic and less likely to have ESRD-HD. Twelve of twenty-seven (44.4%) of ESRD patients had severe disease as opposed to 232/335 (69.3%) of patients not on chronic HD (p=.008).

**Table 1.** Baseline characteristics of hospitalized COVID-19 patients

|  | Entire Cohort<br>(n=362) | Non severe<br>illness (n=130) | Severe illness<br>(n=232) | p<br>value |
| --- | --- | --- | --- | --- |
| Age, years - median (IQR) | 71 (59–82) | 70 (58–78) | 73 (60–84.5) | .1 |
| Gender – Male, n (%) | 200 (55.3) | 68 (52.3) | 132 (56.9) | .4 |
| Race |  |  |  |  |
| Caucasian | 123 (33.9) | 43 (33.1) | 80 (34.5) | .8 |
| African American | 128 (35.4) | 49 (37.7) | 79 (34.1) | .5 |
| Hispanic | 71 (19.6) | 18 (13.9) | 53 (22.9) | **.04** |
| Others | 40 (11.1) | 20 (15.4) | 20 (8.6) | .05 |
| Comorbidities |  |  |  |  |
| Hypertension | 241 (66.6) | 85 (65.4) | 156 (67.2) | .7 |
| Diabetes | 154 (42.5) | 50 (38.5) | 104 (44.8) | .2 |
| Cardiac diseases | 119 (32.9) | 42 (32.3) | 77 (33.2) | .9 |
| Renal disease |  |  |  |  |
| CKD | 41 (11.3) | 12 (9.2) | 29 (12.5) | .4 |
| ESRD-HD | 27 (7.5) | 15 (11.5) | 12 (5.2) | **.03** |
| COPD | 50 (13.8) | 21 (16.2) | 29 (12.5) | .3 |
| Obesity | 123 (33.9) | 41 (31.5) | 82 (35.4) | .5 |
| LOS median (IQR) | 6 (4–10) | 5 (3–7) | 8 (5–12.5) | **<.001** |
| Mortality, n (%) | 149 (41.2) | 0 (0) | (64.2) | **<.001** |

CKD, chronic kidney disease; COPD, chronic obstructive pulmonary disease, ESRD, end-stage renal disease on hemodialysis; IQR, inter quartile range; LOS, length of hospital stay; n, number.

Patients with ESRD-HD had lower odds of severe pneumonia on univariate analysis compared to patients without kidney disease (Table 2). Hispanics had higher age-adjusted odds of severe disease while patients with ESRD-HD had lower odds for the same (Table 3). On multivariable logistic regression, advancing age and Hispanic ethnicity had higher odds of severe disease/pneumonia. In contrast, patients with ESRD-HD [OR 0.4 (0.2–0.9) p=.04] demonstrated lower odds of severe disease compared to patients without kidney disease (Table 3 and Figure 1A).

**Table 2.** Univariate analysis of patient characteristics for severe pneumonia

|  | Odds Ratio (Confidence Interval) | p value |
| --- | --- | --- |
| Age (1-year increments) | 1.01 (0.9 – 1.03) | .08 |
| Male sex (ref Female) | 1.2 (0.8 – 1.9) | .4 |
| Race (ref Caucasian) |  |  |
| African American | 0.9 (0.5 – 1.5) | .6 |
| Hispanic | 1.6 (0.8 – 3.03) | .2 |
| Others | 0.5 (0.3 – 1.1) | .1 |
| Comorbidities |  |  |
| Hypertension | 1.09 (0.7 – 1.7) | .7 |
| Diabetes | 1.3 (0.8 – 2.01) | .2 |
| Cardiac diseases | 1.04 (0.7 – 1.7) | .9 |
| Renal disease (ref no RD) |  |  |
| CKD | 1.3 (0.6 – 2.7) | .5 |
| ESRD-HD | 0.4 (0.2 – 0.9) | **.04** |
| COPD | 0.7 (0.4 – 1.4) | .3 |
| Obesity | 1.2 (0.8 – 1.9) | .5 |

CKD, chronic kidney disease; COPD, chronic obstructive pulmonary disease, ESRD-HD, end-stage renal disease on hemodialysis; RD, renal disease; ref, reference.

**Table 3.** Risk-factors for severe pneumonia

|  | Age-adjusted<br>OR (CI) | p<br>value | Multivariable<br>OR(CI) | p<br>value |
| --- | --- | --- | --- | --- |
| Age (1-year increments) | – | – | 1.03 (1.01 – 1.04) | **.006** |
| Male sex (ref Female) | 1.3 (0.8 – 2.04) | .2 | 1.3 (0.8 – 2.02) | .3 |
| Race (ref Caucasian) |  |  |  |  |
| African American | 0.9 (0.6 – 1.7) | .9 | 0.9 (0.6 – 1.7) | .9 |
| Hispanic | 2.2 (1.1 – 4.5) | **.03** | 2.2 (1.1 – 4.7) | **.04** |
| Others | 0.6 (0.3 – 1.2) | .2 | 0.6 (0.3 – 1.3) | .2 |
| Comorbidities |  |  |  |  |
| Hypertension | 0.9 (0.6 – 1.5) | .8 | 0.9 (0.6 – 1.7) | .9 |
| Diabetes | 1.3 (0.4 – 2.01) | .3 | 1.4 (0.9 – 2.2) | .2 |
| Cardiac diseases | 0.9 (0.6 – 1.5) | .7 | 0.9 (0.5 – 1.4) | .6 |
| Renal disease (ref no RD) |  |  |  |  |
| CKD | 1.2 (0.6 – 2.4) | .7 | 1.1 (0.5 – 2.3) | .8 |
| ESRD-HD | 0.4 (0.2 – 0.9) | **.04** | 0.4 (0.2 – 0.9) | **.04** |
| COPD | 0.7 (0.4 – 1.2) | .2 | 0.7 (0.4 – 1.2) | .2 |
| Obesity | 1.4 (0.9 – 2.2) | .2 | 1.3 (0.8 – 2.2) | .3 |

CI, Confidence interval; CKD, chronic kidney disease; COPD, chronic obstructive pulmonary disease; ESRD-HD, end-stage renal disease on hemodialysis; OR, Odds ratio; RD, renal disease; Ref, reference.

**Figure 1.**
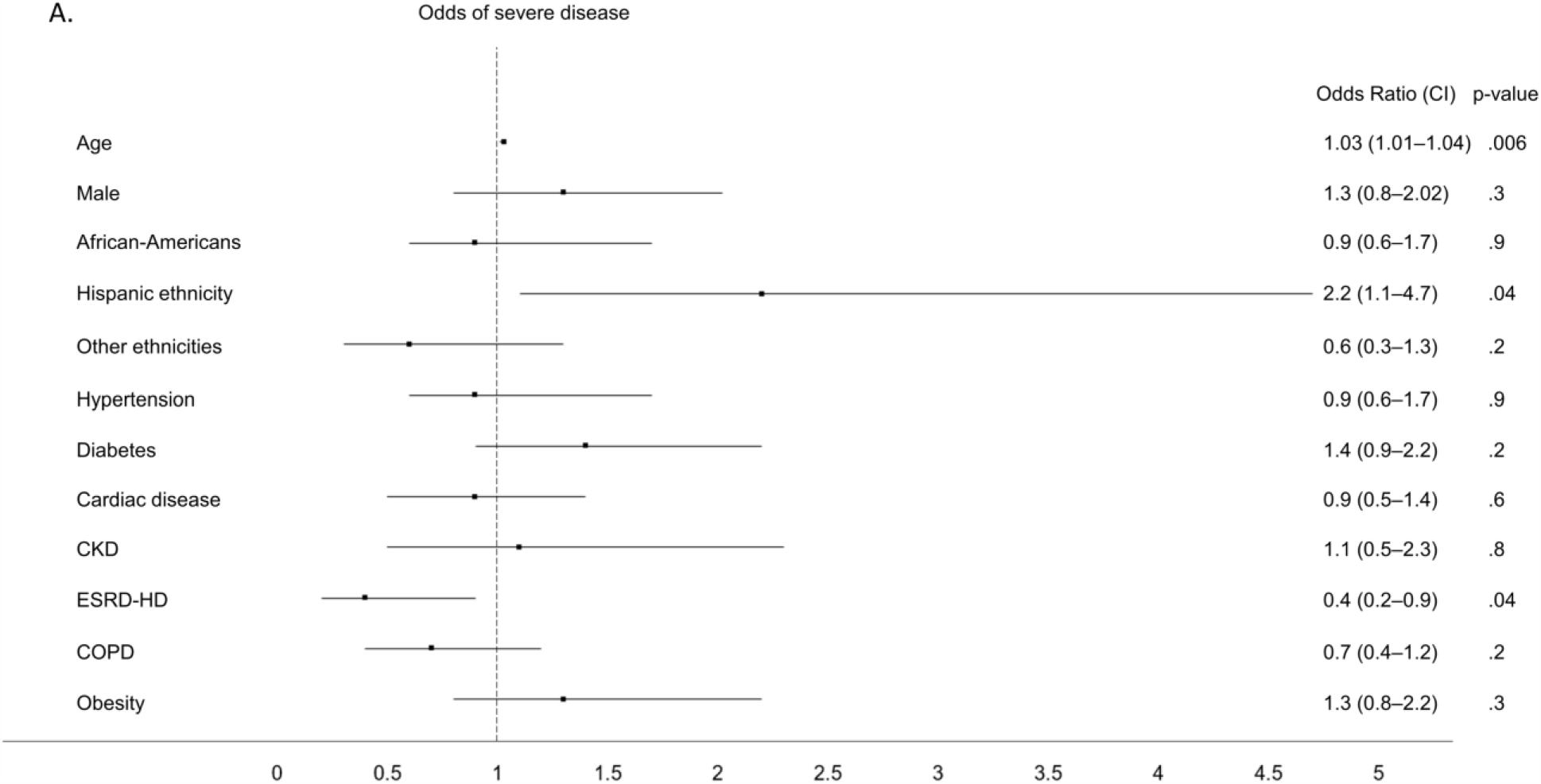
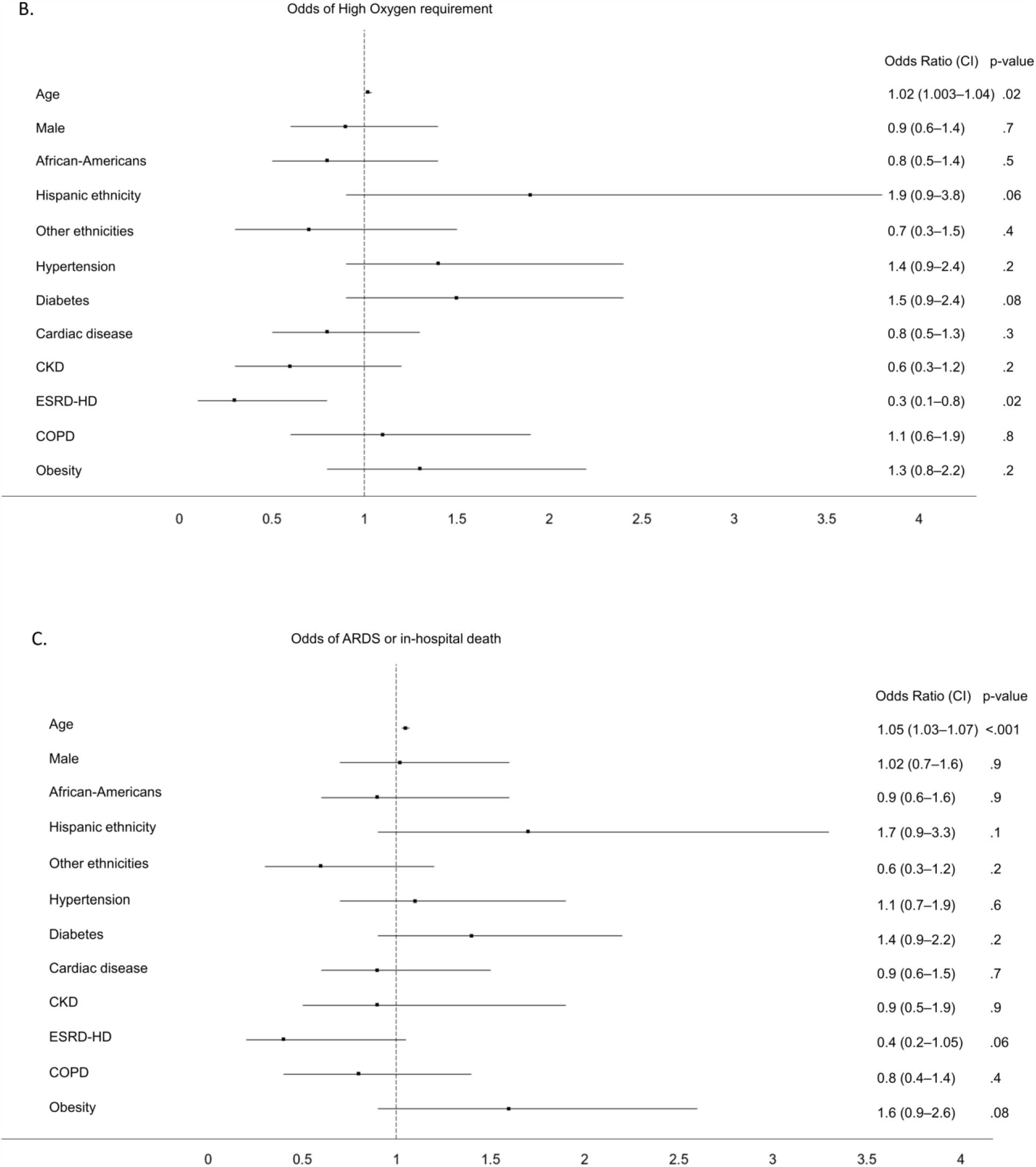
Forest plot showing multivariable odds of severe disease (Figure 1A), odds of the need for high-oxygen support (Figure 1B), and odds of the combined outcome of ARDS and in-hospital mortality (Figure 1C). Abbreviations: ARDS, acute respiratory distress syndrome; CKD, chronic kidney disease; COPD, chronic obstructive pulmonary disease; ESRD-HD, end stage renal disease on hemodialysis.

Additionally, advancing age showed higher odds while ESRD demonstrated lower odds for the secondary outcomes, HiO2 and COAD (Table 4, Table 5, Figure 1B and Figure 1C).

**Table 4.** Risk-factors for high oxygen requirement

|  | Age-adjusted<br>OR (CI) | p<br>value | Multivariable OR<br>(CI) | p<br>value |
| --- | --- | --- | --- | --- |
| Age (1-year increments) | – | – | 1.02 (1.003 – 1.04) | **.02** |
| Male sex (ref Female) | 0.9 (0.6 – 1.4) | 0.8 | 0.9 (0.6 – 1.4) | 0.7 |
| Race (ref Caucasian) |  |  |  |  |
| African American | 0.9 (0.6 – 1.6) | .8 | 0.8 (0.5 – 1.4) | .5 |
| Hispanic | 1.9 (0.9 – 3.6) | .06 | 1.9 (0.9 – 3.8) | .056 |
| Others | 0.8 (0.4 – 1.8) | .6 | 0.7 (0.3 – 1.5) | .4 |
| Comorbidities |  |  |  |  |
| Hypertension | 1.3 (0.8 – 2.2) | .2 | 1.4 (0.9 – 2.4) | .2 |
| Diabetes | 1.5 (0.9 – 2.3) | .06 | 1.5 (0.9 – 2.4) | .08 |
| Cardiac diseases | 0.8 (0.5 – 1.4) | .5 | 0.8 (0.5 – 1.3) | .3 |
| Renal disease (ref no RD) |  |  |  |  |
| CKD | 0.7 (0.4 – 1.4) | 0.4 | 0.6 (0.3 – 1.2) | .2 |
| ESRD-HD | 0.3 (0.1 – 0.9) | .03 | 0.3 (0.1 – 0.8) | **.02** |
| COPD | 1.06 (0.6 – 1.9) | .9 | 1.1 (0.6 – 1.9) | .8 |
| Obesity | 1.4 (0.9 – 2.3) | .1 | 1.3 (0.8 – 2.2) | .2 |

CI, Confidence interval; CKD, chronic kidney disease; COPD, chronic obstructive pulmonary disease; ESRD-HD, end-stage renal disease on hemodialysis; OR, Odds ratio; RD, renal disease; Ref, reference.

**Table 5.** Risk-factors for combined outcome of ARDS or in-hospital death

|  | Age-adjusted<br>OR (CI) | p<br>value | Multivariable OR<br>(CI) | p<br>value |
| --- | --- | --- | --- | --- |
| Age (1-year increments) |  |  | 1.05 (1.03 – 1.07) | **<.001** |
| Male sex (ref Female) | 1.04 (0.7 – 1.6) | .2 | 1.02 (0.7 – 1.6) | .9 |
| Race (ref Caucasian) |  |  |  |  |
| African American | 1.03 (0.6 – 1.7) | .9 | 0.9 (0.6 – 1.6) | .9 |
| Hispanic | 1.7 (0.9 – 3.2) | .1 | 1.7 (0.9 – 3.3) | .1 |
| Others | 0.6 (0.3 – 1.4) | .3 | 0.6 (0.3 – 1.2) | .2 |
| Comorbidities |  |  |  |  |
| Hypertension | 1.1 (0.7 – 1.8) | .6 | 1.1 (0.7 – 1.9) | .6 |
| Diabetes | 1.4 (0.9 – 2.1) | .1 | 1.4 (0.9 – 2.2) | .2 |
| Cardiac diseases | 0.9 (0.6 – 1.6) | .9 | 0.9 (0.6 – 1.5) | .7 |
| Renal disease (ref no RD) |  |  |  |  |
| CKD | 1.1 (0.5 – 2.1) | .9 | 0.9 (0.5 – 1.9) | .9 |
| ESRD-HD | 0.5 (0.2 – 1.1) | .087 | 0.4 (0.2 – 1.05) | .06 |
| COPD | 0.8 (0.4 – 1.4) | .4 | 0.8 (0.4 – 1.4) | .4 |
| Obesity | 1.7 (1.03 – 2.7) | .04 | 1.6 (0.9 – 2.6) | .08 |

CI, Confidence interval; CKD, chronic kidney disease; COPD, chronic obstructive pulmonary disease; ESRD-HD, end-stage renal disease on hemodialysis; OR, Odds ratio; RD, renal disease; Ref, reference.

## Discussion

In this study we present data identifying patient-characteristics that affect illness-severity of people hospitalized with COVID-19. The findings of decreased illness-severity with ESRD in hospitalized COVID-19 patients is novel and needs further investigation. No such association was seen in patient with chronic kidney disease not on HD. This was also observed first-hand by the authors by being on the frontlines in New Rochelle, New York, one of the first COVID-19 epicenters in the US.^5^ Only one other group of researchers have described a lower severity of disease in dialysis patients with COVID-19.^10^ In that study, Ma et al. describe mostly mild clinical symptoms in patients with ESRD and a low likelihood of progression to severe pneumonia in ESRD-HD patients in an outpatient setting. To the best of our knowledge our study is the first to demonstrate a lower illness severity in ESRD-HD patients in the inpatient setting. Of note some studies have mentioned poor outcomes in ESRD-HD patients.^11,12^ However, none of the studies to date have compared outcomes between ESRD-HD patients and patients not on hemodialysis, like we did in the present study.

Additionally, our study unequivocally demonstrated higher illness severity and worse outcomes with advancing age. This is not surprising as by now, advancing age has been established as the most important risk factor for poor outcomes in COVID-19.^13-16^ Male sex and comorbidities (hypertension, diabetes, cardiac disease, obesity, and others) have also been identified as independent risk factors for worse outcomes.^13-15^ In our patient cohort, the odds for severe illness, COAD and HiO2 trended higher among diabetics and obese, although this did not satisfy our pre-determined requirement for statistical significance.

Patients belonging to ethnic and racial minorities are increasingly being identified to be at a higher risk with COVID-19.^17,18^ At first glance it may seem that minorities were not particularly affected in our study, however, a deeper dive into the data would dispel that notion. As per the 2010 US census, the racial/ethnic makeup of New Rochelle was: 48% non-Hispanic Caucasians, 18% African-Americans and 28% Hispanics of any race. Thus, even though we did not see higher odds for poor outcomes among African-American inpatients, they were over-represented among the hospitalized patients when compared to their representation in the community (35% vs 18%). In addition, we found that patients of Hispanic ethnicity had higher odds for severe illness when compared to Caucasians.

Our finding begs the question, what are the pathophysiologic mechanisms behind this association between ESRD-HD and lower illness severity with COVID-19?

In the previous Influenza and novel coronavirus epidemics, severe acute respiratory syndrome (2003) and middle east respiratory syndrome (2012), in addition to virus-induced cytopathic effects, an excessive and dysregulated host immune response played a crucial role in the pathology and mortality.^19,20^ This exaggerated response with supraphysiologic levels of cytokines is referred to as cytokine release syndrome or “cytokine-storm”. Cytokine storm has been described as a major determinant of severity of illness in COVID-19 patients as well.^21-24^ Thus, during viral epidemics, illness severity and outcomes depend on the balance between the beneficial anti-viral effect of the immune response and the harm from an exaggerated inflammatory reaction.

ESRD-HD is simultaneously associated with immune activation leading to systemic inflammation and immune deficiency.^25^ Activation of the constituents of the innate immune system; monocytes, macrophages, and granulocytes, is responsible for ESRD-associated inflammation. On the other hand, immune deficiency in ESRD is caused by the depletion of dendritic cells, memory T cells, B cells and impaired phagocytic function of neutrophils and monocytes.^25^ Thus, one would expect ESRD-HD patients to have a poor antiviral response and an exaggerated inflammatory response with COVID-19, potentially manifesting as poor outcomes in these patients.

An immune-deficient state was indeed demonstrated by Ma et al. in their study.^10^ A significantly lower number of T cells, natural killer cells, and B-cells in ESRD-HD patients were seen compared with patients not on dialysis.^10^ These numbers in ESRD-HD patients with SARS-CoV-2 infection were further decreased. However, paradoxically, the serum levels of inflammatory markers (Interleukin-4, Interleukin-6, and tumor necrosis factor-alpha) were highest in non-HD patients with COVID-19 infection, followed by non-HD patients without COVID-19, and the lowest levels of inflammatory markers were observed in ESRD-HD patients irrespective of COVID-19 infection.

Thus, on one hand, ESRD-HD patients were at a higher risk due to the decreased ability for an effective anti-viral response from an immunodeficient state, on the other hand they benefitted from a lower propensity for inflammatory tissue damage secondary to a dampened cytokine release and a lower likelihood of cytokine storm. Our observation of lower likelihood of severe disease and lesser requirement for oxygen supplementation, suggests that the balance between the benefit from anti-viral immune response and harm from associated inflammatory damage is tilted favorably for individuals with ESRD on HD.

This raises another question, as to why do ESRD-HD patients, who chronically demonstrate an inflammatory state,^26-28^ had significantly lower levels of inflammatory markers in COVID-19 infection compared to patients not on HD?^10^ Perhaps, it is possible that “preconditioning” with this chronic underlying inflammation in ESRD-HD patients attenuates the inflammatory response from an acute insult such as the COVID-19 infection, thereby decreasing the inflammatory damage, illness severity and the need for supportive therapy? This hypothesis is analogous to the well-established “obesity paradox”, in which preconditioning with chronic inflammation in obese patients is thought to contribute to a dampened inflammatory response leading to better outcomes in acute critical illness.^29-30^ Additionally, there is a possibility that the inflammatory cytokines that are generated in response to the acute infection are removed by dialysis membranes during hemodialysis.^31,32^

Another mechanism for better outcomes in chronic-dialysis patients could be related to the renin-angiotensin system. It has been suggested that excessive activation of the Renin-Angiotensin system (RAS) might contribute to progression of COVID-19 related lung injury.^33^ Chronic perturbations of the RAS and intra-renal RAS in ESRD-HD have been described.^34,35^ Modifications in the activation of RAS in response to the acute COVID-19 illness in patients with ESRD-HD might explain the “protective effect” seen in these patients.

Previously described phenomenon of “reverse epidemiology”, the finding that conventional cardiovascular risk factors such as obesity, hypertension, and dyslipidemia are not associated with increased mortality in the hemodialysis population; also point to the fact that ESRD-HD patients are a distinct subgroup and do not respond to risks and insults the same way as general population.^36,37^ From our observation and the findings from the study by Ma et al., it is likely that the altered immune milieu and physiology in patients with end-stage renal disease modifies the inflammatory (cytokine) response to acute COVID-19 illness in these patients, giving them an advantage in the short term. Our findings are novel and of great general medical interest, and should be shared, corroborated and elucidated.

## Conclusions

Patients with ESRD-HD had a milder course of illness with lower likelihood of development of severe pneumonia and lower need for aggressive oxygen supplementation when compared to patients not on chronic dialysis. Our observations suggest that the balance between the benefit from anti-viral immune response and harm from associated inflammatory damage is tilted favorably for individuals with ESRD on HD. This “protective effect” might have a pathophysiologic basis and needs to be further explored. Additionally, age had the greatest impact on illness severity, oxygen requirement and mortality in patients hospitalized with COVID-19. African-Americans were over-represented among the hospitalized COVID-19 patients compared to the community and Hispanics had a more severe illness course compared to Caucasians.

## Strengths and limitations

Limitations include a patient cohort that comprised of only hospitalized patients, from a single hospital and included older individuals and thus, the findings might not be generally applicable. We did not review the effect of medications targeting the RAS (acetyl cholinesterase inhibitors and angiotensin receptor blockers) on illness severity.

Strengths of the study include the accuracy of data that was manually extracted from patient charts, a relatively large cohort and using a standardized model building technique (Lasso) for multivariable analysis.

## Data Availability

The data that support the findings of this study are available from the corresponding author, upon reasonable request.

## Supplementary Materials

Supplementary File (PDF) is available for reference.

